# The Politics of Implementing the COVID-19 Mass Vaccination Programme in Ghana: A Qualitative Exploration Study

**DOI:** 10.64898/2026.07.28.26359091

**Authors:** David E. Akpakli, Rosemond Akosua Nyamedor, Ramadan Haruna, Merri Iddrisu

**Author notes:** Corresponding Author: (DEA).

## Abstract

**Background:** The COVID-19 pandemic disrupted Ghana’s health, economic, and social systems, prompting the government to launch a mass vaccination programme in March 2021, targeting 20 million people by the end of 2022, to reduce transmission and mitigate socio-economic strain. This study explored stakeholders’ perspectives on the politics of the COVID-19 vaccination programme, highlighting implementation challenges, political dynamics, and lessons for future pandemics.

**Methods:** A qualitative, exploratory, descriptive study was conducted between June and September 2021. Fourteen stakeholders from the Ministry of Health, the Ghana Health Service, academia, political leaders, and professional associations were purposively selected and engaged in key informant interviews. Data were analysed using Braun and Clarke’s thematic analysis approach, guided by the Health Policy Triangle and Campos and Reich’s policy implementation framework.

**Results:** Three main themes were generated. First, governance and institutional capacity shaped implementation through issues of procurement transparency, communication gaps, donor dependence, and operational constraints in data systems and logistics. Second, the political economy of vaccine procurement and distribution was influenced by leadership politics, bureaucratic dynamics, fiscal constraints, donor priorities, and interest-group influence. Third, stakeholders highlighted lessons for future pandemic preparedness, including strengthening transparent governance, improving public communication, building local vaccine research and manufacturing capacity, enhancing intersectoral coordination, and establishing robust monitoring and evaluation systems.

**Conclusion:** The rollout of COVID-19 vaccines in Ghana was shaped not only by technical and logistical considerations but also by complex political and governance dynamics. The study highlights how governance structures, political incentives, and interactions among key stakeholders influence policy implementation in resource-constrained settings. Ghana’s experience underscores the importance of transparent governance, effective multistakeholder coordination, sustainable financing mechanisms, and strategic investment in regional vaccine research and manufacturing as critical pillars of pandemic preparedness. Strengthening these institutional and governance dimensions is essential not only for sustaining immunisation programmes but also for building resilient health systems capable of responding effectively to future global health emergencies.

## Introduction

The COVID-19 pandemic posed unprecedented challenges to health systems, governance structures, and socio-economic stability worldwide (1). While mass vaccination was widely promoted as the most effective strategy for controlling transmission and reducing mortality, its implementation was not merely a technical or biomedical process (2,3). Rather, vaccine rollout was deeply embedded in political, institutional, and governance arrangements that shaped decisions about procurement, financing, prioritisation, distribution, and public communication (4,5). In low- and middle-income countries (LMICs), these processes were further complicated by donor dependence, limited fiscal space, infrastructural constraints, and weak accountability mechanisms (5–7).

Globally, governments adopted non-pharmaceutical interventions such as border closures, physical distancing, and mask mandates to curb the spread of COVID-19 (1). Although these measures helped slow transmission, they also generated significant economic and social disruptions. Consequently, mass vaccination emerged as a central pillar of pandemic response. As of August 2025, over 13.64 billion COVID-19 doses had been administered globally, with approximately 67% of the global population completing a primary vaccination series (8). However, coverage remains uneven, reflecting persistent global inequities in access, financing, and supply (2,3). Many African countries, including Ghana, experienced delayed and irregular vaccine deliveries, constrained cold-chain capacity, and fluctuating public demand (9–11).

Ghana was the first African country to receive COVAX vaccines in 2021 and has been applauded for the expanding coverage (12,13). By June 2024, approximately 56% of the population had completed a primary vaccination series, although coverage later stabilised around 42% by August 2025, indicating declining momentum. Despite early political commitment and international support, the rollout encountered persistent challenges, including procurement delays, uneven distribution, vaccine hesitancy, logistical bottlenecks, and misinformation (14,15). These challenges highlight that vaccine implementation outcomes are shaped not only by supply availability but also by governance arrangements, institutional capacity, and political dynamics.

Existing studies in Ghana have primarily explored health system responses, financing, and service delivery during the COVID-19 pandemic (14–17). However, relatively limited attention has been paid to the political and governance dynamics of the COVID-19 vaccination programme, the political and governance processes that influenced vaccine procurement, allocation, and uptake. Understanding these processes is essential, as political leadership, bureaucratic coordination, donor relations, and interest group influence can either facilitate or undermine implementation. Without adequate transparency, accountability, and stakeholder alignment, even well-designed vaccination programmes may fail to achieve equitable and sustainable coverage.

This study addresses that gap by examining stakeholders’ perspectives of the politics of Ghana’s COVID-19 vaccination programme. Specifically, it explores how leadership politics, bureaucratic structures, budgetary processes, donor relations, and beneficiary engagement influenced implementation outcomes.

Several studies have examined COVID-19 vaccine rollout and implementation challenges in African countries, including Nigeria, Kenya, Uganda, and South Africa (10,18–20). These studies have highlighted issues such as supply shortages, health system capacity constraints, vaccine hesitancy, and inequitable access. While this literature provides very useful insights into operational and demand-side barriers, it has largely focused on service delivery and health system performance, with relatively limited attention to the political and governance processes shaping implementation.

In contrast, the present study focuses explicitly on the political economy and governance dimensions of vaccine rollout in Ghana. This study extends existing research beyond descriptive accounts of implementation challenges. It offers a context-specific and theoretically grounded understanding of how political and institutional factors influenced vaccination outcomes, thereby contributing new empirical and analytical insights to the growing literature on pandemic governance in sub-Saharan Africa. The study is guided by Walt and Gilson’s health policy triangle (21) and Campos and Reich (22) policy implementation framework to analyse the roles of stakeholders and contextual elements. While the Health Policy Triangle offers a broad conceptual framework for understanding health policy dynamics, Campos and Reich’s model operationalises political context by identifying key stakeholder groups whose interests and power relations affect implementation.

This study provides a theoretically informed analysis of how political and institutional factors shaped Ghana’s COVID-19 vaccination rollout by integrating these frameworks. It contributes to the health policy and global health literature by demonstrating how governance arrangements, actor coalitions, and financing structures influence policy execution in resource-constrained settings. The findings offer practical lessons for strengthening transparency, coordination, and accountability in future pandemic responses in Ghana and other LMICs.

The objectives set to be achieved in the study include: investigating governance and accountability mechanisms in Ghana’s COVID-19 Vaccination programme, assessing stakeholder perspectives on the political dynamics influencing vaccine procurement and distribution processes, and identifying lessons from the COVID-19 vaccination rollout to inform pandemic preparedness in Ghana and strengthen vaccination programmes in LMICs. This study draws on two complementary frameworks: Walt and Gilson’s Health Policy Triangle (Figure 1) and the Policy Implementation Framework by Campos and Reich (Figure 2). The Health Policy Triangle offers a structured approach to analysing health policy by examining the dynamic interplay among four core elements: actors, context, content, and process (21). Actors in the policy-making process are individuals, groups, organisations, or institutions that participate and have a role in shaping and influencing policy decisions (23). Each actor has distinct roles, interests, and levels of influence in shaping policies (24) and can alter policy content by influencing the process within their professional and social contexts (21).

**Figure 1:**
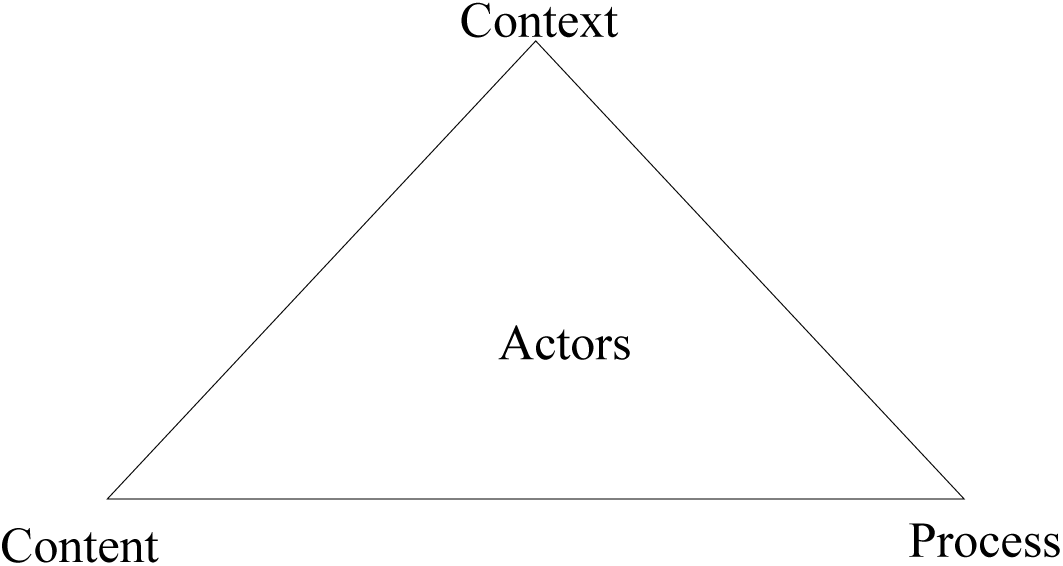
The Health Policy Triangle Source: Constructed by the authors based on (Walt & Gilson, 1994).

**Figure 2:**
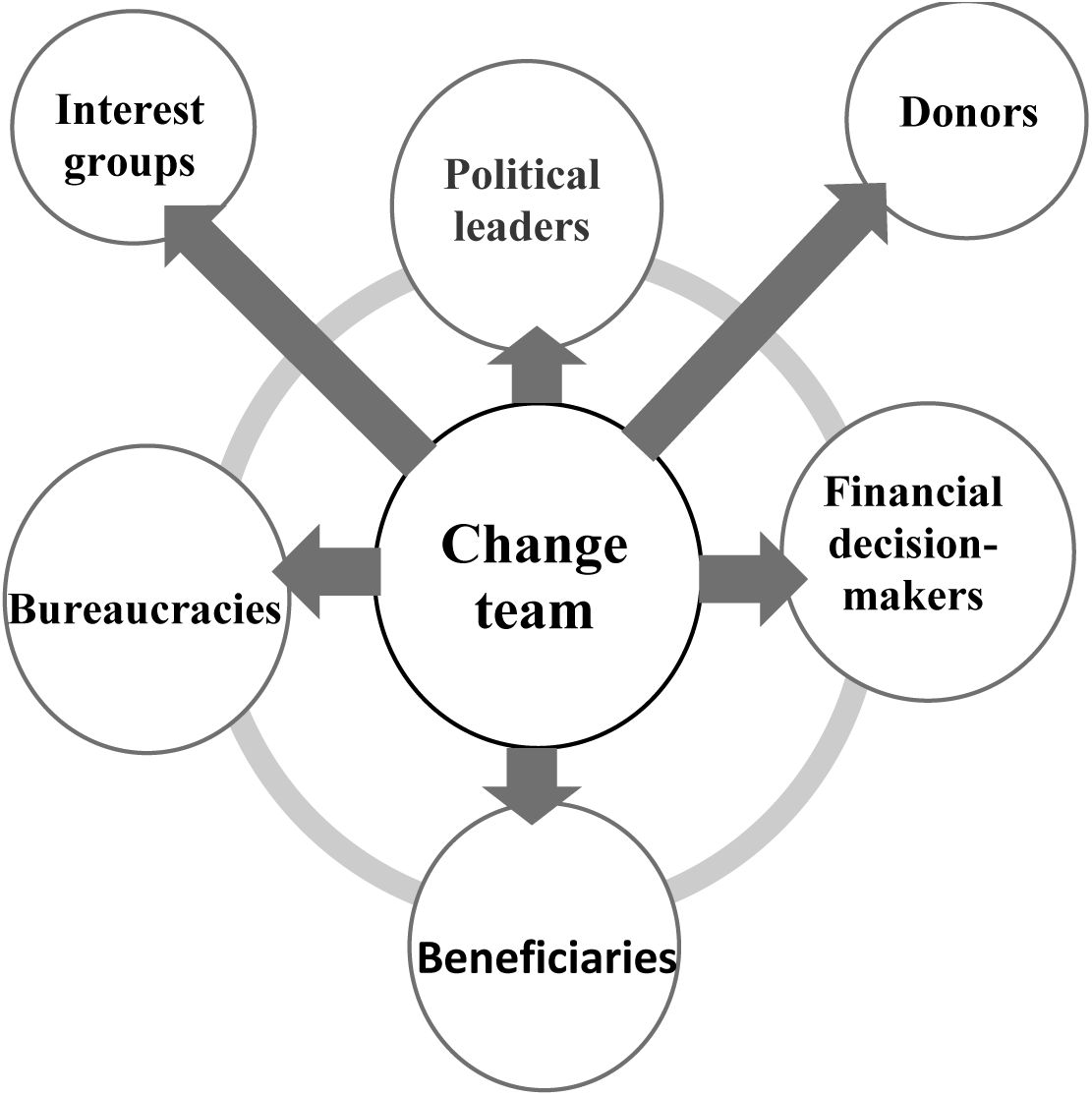
Policy Implementation Stakeholders Source: Adapted from Campos and Reich (2019).

While the policy triangle is a highly helpful tool for analysing health policy research, it offers limited operational guidance for identifying specific stakeholder groups and understanding how they exert influence.

To address this, Kodua et al. (25) linked the actors and context elements of the policy triangle to the politics of the policy process, especially in terms of stakeholder influence. Building on this premise, we use Campos and Reich’s Policy Implementation Framework to operationalise the political context of the COVID-19 vaccination policy process. This framework identifies six stakeholders whose roles, interests, and power must be managed to an enhance effective implementation. These include Political Leaders also known as Leadership Politics; Interest Groups, or Interest Group Politics; Bureaucracies, or Bureaucratic Politics; Financial Decision Makers, or Budget Politics; Beneficiaries, or Beneficiary Politics and External Donors, or Donor Politics (22).

Leadership Politics refers to securing the commitment of political leaders whose vision, expertise and ethical grounding are essential for guiding the processes of policy implementation (22). In Ghana, overcoming vaccine scepticism and logistical barriers required high-level leadership and management capacity. Bureaucratic Politics involve managing frontline bureaucrats, especially within the Ministry of Health and Ghana Health Service and its affiliated organisations (22). Street-level bureaucrats, as described by Lipsky (26), wield discretion that can shape or hinder policy outcomes. Interest Group Politics relates to the influence of professional associations and industry groups such as the Pharmaceutical Manufacturers Association, pharmacists’ professional associations, and medical professional groups, which often try to influence health policy to reduce their losses and increase their gains from the suggested changes (22). Budget Politics concerns decisions about resource allocation, especially during health emergencies (22). The Ministry of Finance’s distribution of public funds across several sectors can constrain the Ministry of Health’s ability to implement vaccination strategies effectively. Donor Politics examines the role of international donors and agencies, whose funding, technical expertise, and priorities can significantly influence national health policy processes, sometimes challenging local ownership and priorities (22). Beneficiary Politics focuses on engaging the public as end users of health policy (22). Beneficiaries’ trust, awareness, and active participation are crucial for successful uptake, but can be undermined by misinformation, apathy, and conflicting messages from stakeholders (22).

However, stakeholder categories may overlap. Bureaucrats may also be beneficiaries; leaders can be part of interest groups; and opposition political leaders may act as pressure groups. For example, the Pharmaceutical Society of Ghana and the Ghana Medical Association include members who serve as bureaucrats in various healthcare facilities across the country. Recognising and managing these overlaps are essential for understanding the political governance challenges encountered during Ghana’s COVID-19 vaccination programme.

## Methods

### Study Design

An exploratory, descriptive, qualitative approach was used to investigate the phenomenon, as little is known about it, and the researchers were interested in exploring and describing stakeholders’ perceptions of Ghana’s COVID-19 vaccination programme. We chose this approach to capture the nuanced insights into governance, political dynamics, and operational challenges (27).

#### Population

The study targeted all key stakeholders involved in the procurement and distribution of the COVID-19 vaccine in Ghana.

#### Study Setting

This study was conducted in the Greater Accra Region of Ghana during the COVID-19 vaccination rollout. The period of interest covered the programme’s initiation and subsequent phases, capturing significant developments in procurement, distribution, and public engagement.

#### Sampling Participants

A purposive sampling technique was used to recruit participants with direct involvement in or oversight of Ghana’s COVID-19 vaccination programme. Participants were selected based on their institutional roles, technical expertise, and engagement in vaccine procurement, coordination, implementation, and community mobilisation. This approach ensured the inclusion of diverse perspectives from key stakeholder groups, including government officials, health professionals, development partners, and civil society actors.

The final sample comprised 14 participants. Sample size was guided by the principle of information power, which suggests that fewer participants may be sufficient when the study has a focused aim, participants are highly relevant to the research questions, and the data collected are rich and of high quality (28). In this study, information power was assessed based on five criteria: (1) a focused study aim centred on governance and political dynamics; (2) participants’ specialised knowledge and direct involvement in vaccine implementation; (3) the use of established theoretical frameworks to guide analysis; (4) in-depth, semi-structured interviews that generated rich and detailed data; and (5) strong dialogue quality between researchers and participants.

Data collection continued until sufficient informational redundancy was achieved, whereby no substantively new themes were emerging from subsequent interviews. At this point, the research team determined that the dataset provided adequate depth and breadth to address the study objectives.

Data were collected between June and September 2021 through semi-structured key informant interviews. Interviews were conducted either in person or virtually, depending on participants’ availability and prevailing public health guidelines. Each interview lasted approximately 45 to 60 minutes and was audio-recorded with participants’ consent.

### Data collection tool and procedure

The researchers developed a semi-structured interview guide based on the study’s objectives, which were carved from the Health Policy Triangle and Campos and Reich’s Policy Implementation Frameworks (22,27). The guiding questions were divided into two parts: demographic characteristics constituted one, and the main guiding question formed the second part. After ethics approval, the first author approached the targeted participants with the information sheet outlining the rationale for the study and sought their permission to be interviewed. The questions were piloted to ascertain their completeness before the main data collection. No changes were made to the questions after the pilot. The majority of the interviews were conducted face-to-face at participants’ offices, except for one conducted via Zoom. With participants’ consent, interviews were audio-recorded and complemented by note-taking. The discussions explored governance and accountability mechanisms, political influences on procurement and distribution, and lessons for future pandemics.

### Data Analysis

Data were analysed following Braun and Clarke’s (29) six-phase thematic analysis framework: (1) familiarisation with the data, (2) generating initial codes, (3) searching for themes, (4) reviewing themes, (5) defining and naming themes, and (6) producing the report. Interview transcripts were analysed using thematic analysis. Two researchers independently coded all 14 transcripts using Microsoft Excel to organise and manage the data. Each transcript was first read in full to ensure familiarisation with the content. The transcripts were then segmented into meaningful units and entered into Excel, where columns were used to record initial codes, analytic notes, and emerging categories. Initial codes were generated inductively from recurring ideas and patterns relevant to the study objectives. During the coding process, a shared codebook was developed and refined iteratively as new codes emerged and existing codes were clarified. After independent coding, the researchers compared coded transcripts to identify similarities and differences in code application. Coding disagreements were discussed in a series of analytic meetings, during which the researchers revisited the original transcript excerpts and reached consensus through discussion. The Principal Investigator provided oversight to ensure consistency and analytical rigour. Related codes were grouped into broader themes, and overlapping codes were refined or merged to strengthen conceptual clarity through iterative discussions within the research team. To enhance the credibility of the findings, interpretations were triangulated across the research team through collective review of coded excerpts and emerging themes. The final themes were then synthesised and organised to capture the political dynamics surrounding COVID-19 vaccine procurement and distribution.

#### Rigour

Several strategies were adopted to ensure the rigour and trustworthiness of this qualitative study. Credibility was enhanced by triangulating perspectives across diverse stakeholder groups, including policymakers, programme managers, frontline health workers, professional associations, and political actors. Two members of the research team independently coded the transcripts using Braun and Clarke’s six-phase thematic analysis approach, and discrepancies were resolved through discussion until a consensus was reached. This iterative process supported inter-coder reliability and consistency in theme development. Informal member checking was also undertaken, with selected participants consulted to confirm the accuracy of interpretations. To ensure dependability, the research team maintained a clear audit trail, documenting coding decisions, thematic refinements, and analytical memos throughout the process. The themes and sub-themes are included in the manuscript to allow transparency and replication of the analytic approach. While the sample size was relatively small and concentrated in Greater Accra, the use of rich, contextually grounded narratives enables readers to assess the applicability of the findings to other contexts.

#### COREQ compliance

This study was conducted in line with the Consolidated Criteria for Reporting Qualitative Research (COREQ). Interviews were conducted by the Corresponding Author, a researcher in public health, who had no prior personal relationship with participants. Participants were informed of the researcher’s academic background and the purpose of the study before providing consent. Fourteen key informant interviews were conducted until saturation was reached. Interviews were audio-recorded with permission, transcribed verbatim, and anonymised using unique identifiers. Transcripts were not returned to participants for their review and comment. Coding and analysis were performed manually in Microsoft Excel, applying Walt and Gilson’s policy triangle and Campos and Reich’s framework to guide interpretation.

### Ethical Considerations

Ethical approval was obtained from the Dodowa Health Research Centre Institutional Review Board (DHRCIRB/051/05/21). All participants provided written consent, and the confidentiality of the data was guaranteed. The study adhered to the principles outlined in the Declaration of Helsinki.

#### Author Reflexivity and Positionality

In line with best practices for transparency in qualitative research, the authors reflected on how their professional backgrounds and positions might have influenced the design, data collection, analysis, and interpretation of findings.

The research team comprised members with complementary expertise in public health research, health systems management, political and governance studies, and academic scholarship. Several team members held professional roles within health institutions and research organisations, positioning them as partial insiders in the policy environment under study. This facilitated access to key informants and enhanced participants’ willingness to engage. However, it may also have reinforced power asymmetries and encouraged socially desirable responses, selective disclosure, or cautious framing of politically sensitive issues.

Team members involved in fieldwork and data analysis also possessed prior experience working with some participants and institutions. While this familiarity supported rapport and trust-building, it carried the risk of confirmation bias and the normalisation of institutional practices. In addition, researchers’ academic and professional status may have influenced how participants perceived the study, potentially shaping expectations about confidentiality, reporting, and policy relevance.

To mitigate these potential influences, the team employed multiple reflexive and methodological strategies. These included the use of open-ended and neutral interview techniques, explicit assurances of confidentiality and independence, maintenance of reflexive journals, and documentation of analytic decisions. Regular peer debriefing sessions and collaborative analysis meetings were conducted to interrogate assumptions, challenge dominant interpretations, and enhance critical reflexivity. Where possible, divergent perspectives were actively sought and incorporated into theme development.

Rather than seeking to eliminate subjectivity, the authors acknowledged their positionality as integral to knowledge production within an interpretivist-constructivist paradigm. Through sustained reflexive practice, transparent documentation, and collective sense-making, the team sought to manage power dynamics and potential biases while strengthening the credibility, dependability, and trustworthiness of the findings.

## Results

A total of 14 key informants participated in the study, representing multiple stakeholder groups involved in the COVID-19 vaccination programme in Ghana, including government ministries, health service implementers, professional associations, academic institutions, political actors, civil society and religious organisations. Details of participants recruited are provided in table 1 below.

**Table 1.**
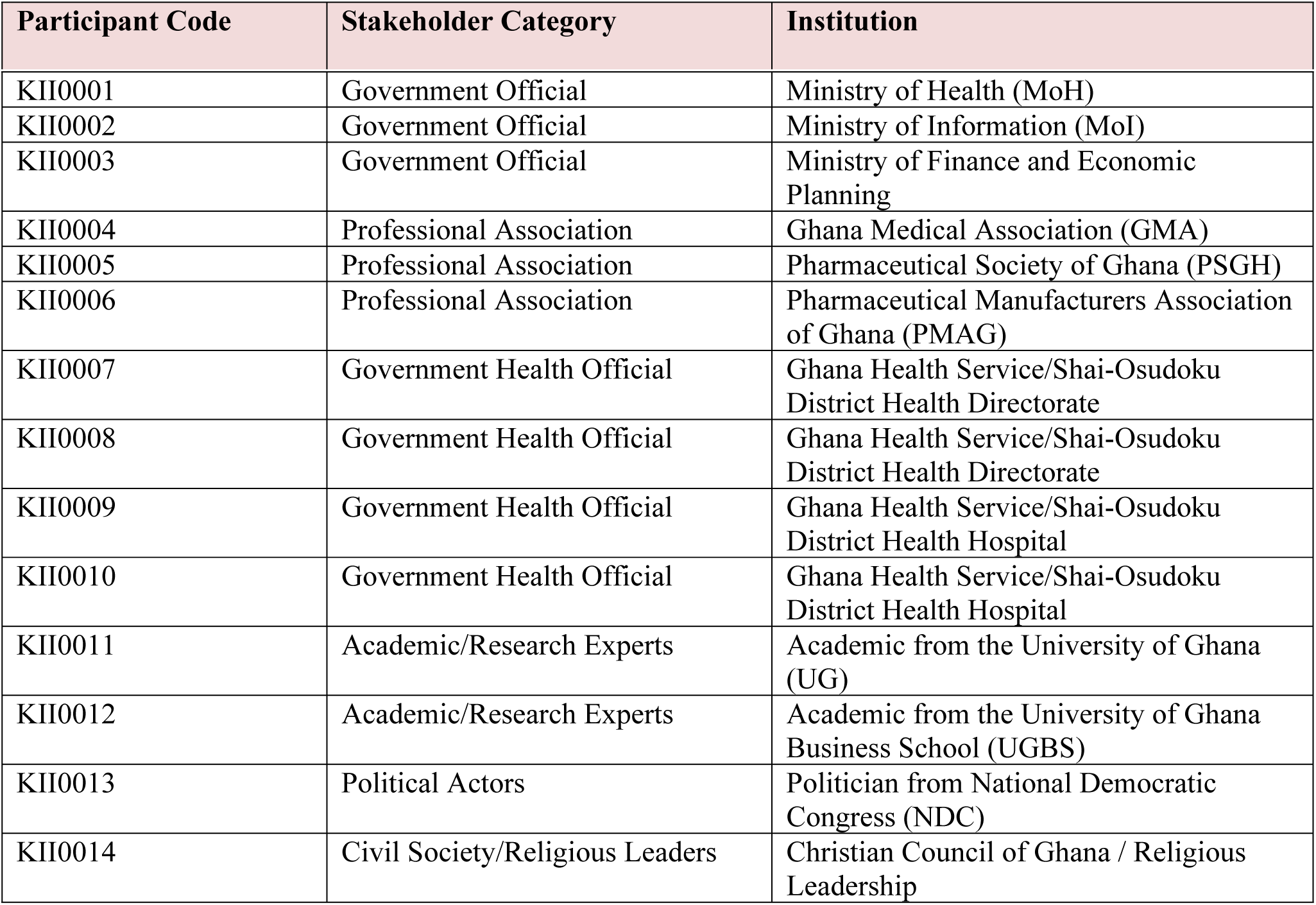
Characteristics of Study Participants (n = 14)

### Themes and subthemes

The Thematic analysis yielded three overarching themes with nine interrelated subthemes, as summarised in Table 2 below.

**Table 2.**
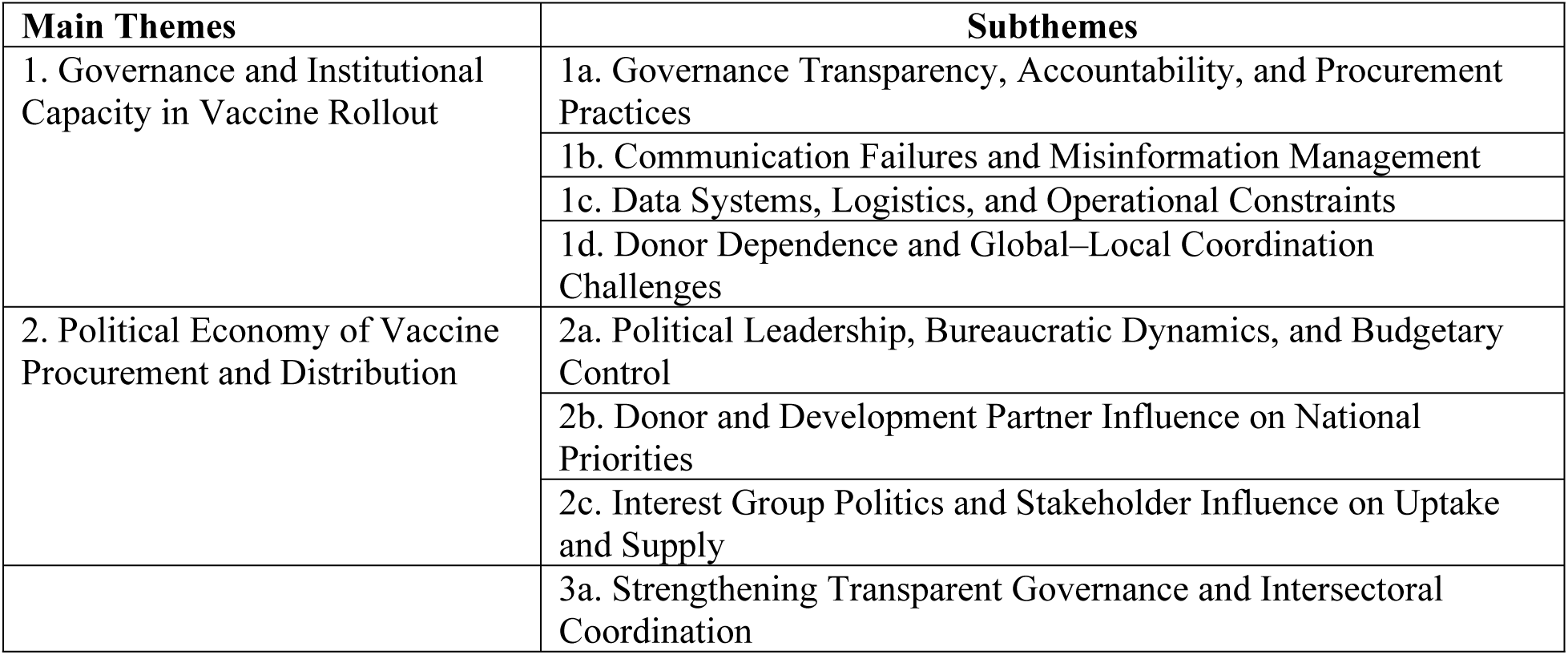
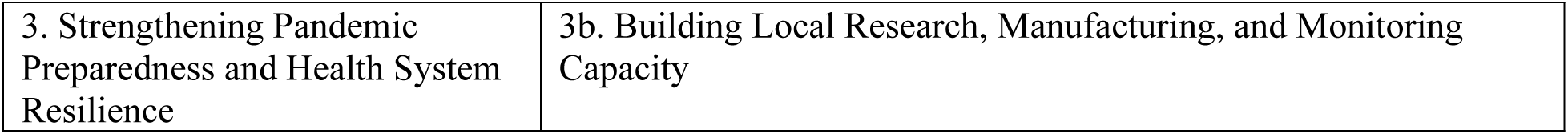
Main Themes and Subthemes.

### 1. Governance and Institutional Capacity in Vaccine Rollout

Stakeholders emphasised that governance and institutional capacity were critical determinants of the success or failure of Ghana’s COVID-19 vaccination programme. While early and visible political commitment proved essential in securing initial vaccine supplies and mobilising public campaigns, weaknesses in procurement transparency, communication strategy, coordination, and operational systems constrained effective implementation. The analysis identified four interrelated dimensions: governance transparency and accountability; communication and misinformation management; donor dependence and global coordination; and operational constraints in data and logistics.

#### 1a. Governance Transparency, Accountability, and Procurement Practices

Concerns about transparency in procurement and financial management featured prominently across stakeholder groups. Participants reported that limited visibility into procurement decisions and fund disbursement processes weakened public trust and fuelled perceptions of politicisation.

Several stakeholders suggested that procurement processes were insufficiently transparent and may have been influenced by personal or political interests. As one frontline healthcare worker remarked:

> *“I will always procure where I would benefit as an individual or where in future, I would get something in return… Recently, I even heard news that one minister in charge had an interest in it and therefore added his share to it; that is why the vaccines were so expensive*.” (KII0010).

Such perceptions, regardless of their factual basis, eroded confidence in the fairness and integrity of the process.

Concerns also extended beyond procurement to financial accountability at subnational levels. A district-level actor noted that funds allocated for COVID-19 activities were not always transparently communicated to implementation teams, creating uncertainty about resource distribution and entitlements. Similarly, questions were raised about the criteria used for prioritising early vaccine recipients. One academic stakeholder observed that the selection process for initial vaccine allocation “was not clearly communicated, leading to public grumbling and suspicion.”

> *“I don’t think the process and strategies involved in the distribution and administration was transparent. We were just sitting there when the announcement came… The committee had some criteria for selecting people, but it wasn’t clear. There was a lot of grumbling… “* (KII0011).

Other stakeholders echoed similar concerns, arguing that clearer financial reporting and more proactive disclosure could have strengthened legitimacy. The absence of routine, publicly accessible reporting mechanisms contributed to speculation about elite capture, particularly during the distribution of early vaccine consignments. Collectively, these perceptions underscore how transparency deficits can undermine programme credibility, even in contexts where political commitment is visible.

> *“With regards to the funds that were received at the directorate, I think that how the funds were disbursed was a bit questionable. The government itself should be a little bit more transparent in what they tell us and also a little bit more forthcoming on information that is necessary.”* (KII0013)
>
> *“It seems as if the information on the vaccination is somewhere, but it’s not known to the public.. …they cannot account for it. It was not transparent… There is no accountability system that reports the day-to-day account of what went in, what was disbursed and others.”* (KII0011).

#### 1b. Communication Failures and Misinformation Management

Participants widely agreed that delayed and fragmented risk communication allowed misinformation to proliferate. Across health, political, and civil society actors, there was a shared view that proactive engagement with communities began too late, leaving space for conspiracy narratives to gain traction:

> *“Some bad people are saying that the vaccine is the mark of the beast* (*666*) *and other false information is also going on. “* (KII0001).
>
> *“We waited for some people to completely destroy the importance of vaccines in our lives through social media. Even some of our big-time media personnel went on the radio and said all sorts of things about the vaccines.”* (KII0004).

A professional association representative reflected that vaccine education “did not start early enough,” enabling social media commentary and influential public figures to shape narratives before official messaging was fully mobilised. Religious and informal opinion leaders were frequently cited as powerful actors in shaping perceptions, sometimes more influential than formal health authorities:

> *“So far, I think, I will tell you candidly, we did not start vaccine education early. We could not start the conversation early. We should have started the discussions on vaccines earlier… There are many people in the country who will not listen to a Minister of Health, but they will listen to a pastor who does not even have knowledge about health issues…”* (KII0004).

Stakeholders acknowledged that misinformation, ranging from religious interpretations to conspiracy theories, circulated widely and was not countered swiftly. One senior official noted that managing “bad press” and misleading social media narratives required stronger coordination and earlier engagement. Participants argued that insufficient early education contributed to hesitancy and slowed uptake, particularly during the rollout of the first doses:

> *“Education was not well done. A lot of people were not happy that they had to take the vaccine. They did not take the first dose of AstraZeneca because they heard all the conspiracy theories on social media, but the government had not spent enough time educating people on what is important.”* (KII0013).

These findings illustrate how communication gaps intersect with governance challenges, reinforcing mistrust and weakening early vaccination momentum.

#### 1c. Donor Dependence and Global-Local Coordination Challenges

Stakeholders highlighted that Ghana’s heavy reliance on donor-supported mechanisms, particularly COVAX, significantly shaped implementation dynamics. Delays and unpredictability in vaccine deliveries disrupted rollout schedules and complicated planning at national and district levels:

> “*Ghana does not have enough vaccines… A good number of people have not been vaccinated; only 2 million out of 30 million had 2 shots because the country was heavily depending on donor support. With all, especially the Sputnik, when we could not get enough of the vaccines from the COVAX… which people thought was less effective.”* (KII0013).

Participants noted that inconsistent supply made it difficult to complete vaccination cycles, with some individuals unable to access second doses within recommended timeframes. As one district-level implementer explained, delivery uncertainties created interruptions in continuity of care:

> *“Sometimes, you can’t even tell when the vaccines will arrive… Some groups of people who took the first vaccine could not access the second vaccine up to date. The time they are supposed to take the second dose of the vaccine has elapsed. This means these people have to start everything all over again..*. (KII0007)

*“Another problem is inconsistency; the second dose is not yet in for those who took the first to go for it.”* (KII0012).

Beyond immediate supply issues, participants raised concerns about repeated political commitments to establish local vaccine manufacturing capacity. Some described these announcements as aspirational but insufficiently institutionalised, reinforcing perceptions of structural dependency on external actors:

> “*Time after time, the president keeps giving us promises that Ghana is doing its best to contact pharmaceutical manufacturers to see whether they can actually start local production of vaccines… all are political talks*” (KII0012).

These accounts underscore how global inequities and donor dependence constrained national autonomy in vaccine planning and implementation.

#### 1d. Data Systems, Logistics and Operational Constraints

Operational bottlenecks, especially in data management and logistics, further constrained implementation. Frontline actors described challenges with digital registration systems, noting that inadequate infrastructure and technical glitches slowed data entry and created administrative backlogs:

> “*At a point in time, we did not really have what we needed to digitalise the vaccination process in such a way that we can avoid people taking advantage of the system.“* (KII0001).
>
> “*We had challenges with the app as well, so those who were in the field, when they were doing the entries and all that, encountered challenges, so what they had to do was some of us entered it manually at the same time, trying to see if we could use the vaccine app.”* (KII0009).

Some participants acknowledged that digital systems were not fully operational at the outset, limiting real-time tracking and increasing vulnerability to system inefficiencies. Field implementers reported difficulties with the vaccination application, often resorting to manual documentation when electronic tools malfunctioned. As one academic stakeholder observed, while the vaccination process itself was relatively efficient, recording and documentation became the bottleneck:

> *“The bottleneck is using electronic thing to capture data, which caused delays in certain places… The process of jabbing was short, but the recording was the problem”.* (KII0011).

Logistical challenges also extended to cold chain management and vaccine shelf life. Participants described instances where consignments arrived with limited time before expiry, placing pressure on rapid distribution and, in some cases, resulting in wastage. Uneven distribution across districts further compounded these inefficiencies:

> “*Do you know the vaccines that came were close to the expiration date? I learnt some even got expired, so next time if they are going for vaccines, they should look for vaccines that had prolonged expiry date, not vaccines that they would bring on Monday and by Friday it’s expired, so it means if you don’t vaccinate within 5 days, you can’t vaccinate any longer.”* (KII0010).
>
> *“…there were certain areas where the vaccines… some even got expired and not fit for use”* (KII0005).

These operational constraints reveal how governance challenges were not only political but also technical and infrastructural, highlighting the interconnectedness of institutional capacity and policy implementation outcomes.

### 2. Political Economy of Vaccine Procurement and Distribution

Ghana’s initial vaccine doses were received through the COVAX facility, followed by additional supplies obtained through bilateral donations and government procurement arrangements. Findings from the study revealed that the procurement and distribution of COVID-19 vaccines in Ghana were significantly shaped by political negotiations and the wider governance environment. Participants consistently described how political leadership, bureaucratic processes, fiscal decisions, donor involvement, and the actions of interest groups influenced the course of implementation. These perspectives closely mirrored the stakeholder categories described by Campos and Reich, underscoring the role of politics in policy execution. Three interrelated dimensions emerged: (1) political leadership, bureaucratic dynamics, and budgetary control; (2) donor and development partner influence; and (3) interest group politics shaping uptake and sustainability.

#### 2a. Political Leadership, Bureaucratic Dynamics, and Budgetary Control

Political leadership played a central role in vaccine procurement decisions, including choices regarding type, sourcing, pricing, and the use of intermediaries. Participants noted that procurement decisions were highly centralised, with limited transparency regarding pricing structures and negotiation processes. As one senior government official observed, key political questions revolved around where the vaccines were procured from, who negotiated them, and whether intermediaries influenced pricing:

> *“One key issue in the politics of this process is the procurement of vaccines. Where they have been procured from, what type of vaccine, who is procuring the vaccine, the middlemen, and how much the vaccine is being procured? Are we buying it at the normal price, or because we are moving into middlemen, the price is being doubled?”* (KII0001).

Beyond procurement, political authority extended into vaccine distribution at the district level. Some participants described instances in which vaccine access was perceived to be influenced by local political considerations. A frontline officer reported that vaccination decisions in their district were subject to political authorisation, creating perceptions of partisan filtering:

> *“Not everyone is getting access to the vaccines. In my district, for example, the vaccination exercise has been politicised. I was told the district head has given a command that until he authorises to give the vaccine to a particular person, it should not be administered to them. Therefore, I was not given because I do not belong to his party.* (KII0008).

While such accounts may not reflect universal practice, they underscore how politicisation narratives can undermine trust in public health programmes.

Participants also noted that vaccine politics operated across scales, from global negotiations to local implementation. Competing interests, ideologies, and institutional actors influenced procurement pathways and distribution decisions. As one participant reflected:

> *“So, the politics of the COVID-19 vaccine is from the global stage to the local level. It is important that we look at it from the perspective that there are various interests, and various beliefs, various people who have various say on vaccines and influences on the vaccines and then the vaccination process, it is diverse, and it is different and varied.* (KII0006).

Bureaucratic dynamics further shaped programme execution. Although the Ministry of Health provided policy direction, operational responsibility rested largely with implementing agencies. Participants described friction between planning and execution, particularly given that existing immunisation structures were historically designed for childhood vaccination rather than adult mass campaigns. One stakeholder noted that while extensive stakeholder engagement occurred during planning, implementation did not fully match the ambition of the plans:

> *“Implementation was not well done, although it was well planned. For instance, the community pharmacies were going to be used as vaccination points; pharmacists were going to be involved in one way or the other (the private pharmacists because of the skill). But when it came to the time, these things were not implemented. They were just implemented partially…”* (KII0005).

Operational discretion also influenced which planned interventions were realised. For example, proposals to integrate community pharmacies into vaccination delivery were reportedly only partially implemented. Participants suggested that unclear role definitions and overlapping institutional mandates contributed to inefficiencies, duplication, and uneven allocation across districts:

> *“I think that there were certain areas where the vaccines that were taken were inadequate. There were others where the vaccines were more than what was taken there, and transferring from one area to another, there were temperature issues… which some even got expired and not fit for use.”* (KII0005).
>
> *“…each of the key stakeholders must be assigned a specific role to play and must be clearly defined… we must have a laid out framework for the disbursement and distribution of the vaccine* (KII0012).

Budget politics compounded these challenges. Although funds were mobilised for procurement, participants reported that financing for deployment, including logistics, outreach, and digital systems, lagged behind. A senior health official acknowledged that “funding for deployment” represented a major constraint. Limited and delayed disbursement particularly affected rural districts, where basic operational resources were already stretched:

> *“…the major challenge we have has to do with funding for deployment of vaccines…”* (KII0002)
>
> *“The challenge is, during the initial vaccination, four rural districts, including ours, were sidelined…sometimes, getting water to drink is even a challenge.”* (KII0009).

Financial constraints also slowed digitalisation efforts and monitoring systems, reinforcing inefficiencies in data capture and coordination. Without adequate funding, robust electronic systems could not be fully operationalised at critical stages of rollout.

> *“At a point in time, we did not really have what we needed to digitalise the vaccination process in such a way that we can avoid people taking advantage of the system.”* (KII0002)
>
> *“The key bottleneck is, we don’t have the vaccines and then the funds to deploy the vaccines.”* (KII0004)

Collectively, these accounts demonstrate how political authority, bureaucratic discretion, and fiscal limitations interacted to shape implementation outcomes.

#### 2b. Donor and Development Partner Influence on National Priorities

Donor support was widely recognised as essential to Ghana’s COVID-19 vaccination programme; however, participants highlighted tensions between external priorities and national needs. Ghana’s reliance on global supply chains and donor mechanisms exposed the country to geopolitical dynamics beyond its control:

> *“Because of the international body politics, it is difficult to get the vaccines.”* (KII0005).
>
> “*We are now losing trust in our developing partners, because of the politics they are playing with the vaccine*.” (KII0011).

Stakeholders described how international politics constrained access to vaccines, especially during the early stages of global distribution. One stakeholder observed that “because of international politics, access became difficult,” while another noted declining confidence in development partners amid perceived inequities in allocation. Participants pointed out that high-income countries secured large advance orders, limiting availability for countries dependent on pooled mechanisms such as COVAX:

> “*There was anxiety in town because everybody was waiting… It is because we are also depending on the developed countries, and unfortunately, in places where we could have had it, like India, the pandemic was so severe, and they have refused to export their vaccines*.” (KII0001).

Reliance on donor-supported supply chains created uncertainty and delays. When exporting countries experienced domestic surges, vaccine shipments were postponed, disrupting local rollout schedules. These disruptions reinforced perceptions of structural vulnerability and highlighted the asymmetry in global health governance:

> *“… procurement of vaccines really has, in the part of our world, been quite challenging. Where initially, none of the producing companies or countries was ready to sell because Europe and America have placed orders”* (KII0003).

Thus, while donor engagement was indispensable, it simultaneously limited national autonomy in procurement timelines and strategic planning.

#### 2c. Interest Group Politics and Stakeholder Influence on Uptake and Sustainability

Interest groups played significant roles in shaping both vaccine uptake and long-term sustainability discussions. Professional associations and pharmaceutical stakeholders advocated for stronger government investment in domestic manufacturing capacity, arguing that procurement guarantees and financial support would reduce future dependency on external suppliers. One participant emphasised that government commitment to purchasing locally manufactured vaccines would be essential to stimulate sustainable production:

> *“Government needs to support pharmaceutical manufacturers financially to produce affordable vaccines… The government must guarantee Ghanaian pharmaceutical manufacturers that their vaccines would be bought… Ghana can do what the National Institute of India is doing… putting structures in place to ensure that pharmaceutical manufacturers can also have the licence to manufacture.”* (KII0004).

Religious leaders emerged as influential actors in shaping public attitudes. Participants described how religious authority extended beyond spiritual guidance into health decision-making, sometimes superseding medical expertise. One stakeholder recounted that even highly trained professionals sought guidance from faith leaders regarding vaccination decisions, illustrating the depth of trust vested in religious institutions:

> *“Let me tell you, even a medical doctor, when the vaccine thing came, was asking me, “pastor, should I go for it?” This should tell you the kind of authority religious leaders have in the community. …medical doctor will still seek advice from a pastor whether he should go for a vaccine. I mean, it doesn’t sound right It’s the reality because of the belief people have in God and in the men and women of God,”*(KII0014).

At the same time, certain doctrinal positions were perceived to discourage engagement with biomedical interventions. Some participants noted that specific religious teachings promoted scepticism toward vaccination, contributing to hesitancy within particular communities:

> *“Culturally, some religions tend to resist these orthodox medicines. We know of some religions which preach that their people should not go to the hospital, and their people, including children, should not be vaccinated. So, these are also challenges which would add up to the politics at the local front*.” (KII0004).

These findings highlight how non-state actors, including faith-based leaders and professional associations, influenced both demand-side dynamics and strategic discussions about future supply resilience.

### 3. Lessons for Future Pandemics

Participants drew on their COVID-19 vaccination experience to identify a focused set of lessons for strengthening Ghana’s preparedness. Central to these were the need for transparent governance and clear accountability in procurement and finance to rebuild trust; sustained and adequately public communication to counter misinformation; strategic investment in local research and vaccine manufacturing to reduce external dependence; stronger intersectoral coordination that integrates ministries, private providers, community health structures and trusted local leaders for faster, more equitable rollout; and robust monitoring and evaluation systems to ensure performance tracking, and corrective action. These reflections generated five interconnected subthemes: strengthening transparent governance, strengthening public communication and education, building local research and vaccine production capacity, enhancing intersectoral coordination and leveraging existing structures and robust monitoring and evaluation systems.

#### 3a. Strengthening Transparent Governance

Participants emphasised that strengthening transparent governance is central to Ghana’s preparedness for future pandemics. They argued that opaque procurement and unclear roles among institutions had fuelled mistrust during the COVID-19 vaccination programme, and stressed that future responses must prioritise accountability at every stage. For many, transparency was an administrative requirement and the foundation for building public confidence and preventing controversies. One academic proposed that the establishment of independent institutions dedicated to managing emergency resources could help safeguard credibility:

> *“Government should set up an independent institute, channel these resources to them, make them accountable state institutions and ensure that expenses and policy tools of these institutions are transparent.”* (KII0011).

Frontline actors similarly stressed the importance of openness in financial disbursement, arguing that community members should be able to track how funds are allocated and spent:

> *“I think we should be transparent about the funds that have been released to the various sectors. It should come on air so that people at the grassroots would know about it. Government should be transparent, accountable and should distribute the resources equitably.”* (KII0008).
>
> *“I believe that when there’s transparency, fairness, and strong advocacy, everyone is on the same page. But sometimes, people from higher levels come in thinking they know everything. They design plans without involving us, and expect us to implement them*.
>
> *…funds are not always disbursed in ways that directly support COVID prevention. Sometimes, they’re used for other purposes.”* (KII0010).

Officials from the Ministry of Health also reinforced this view, calling for clear frameworks that embed accountability into all stages of procurement and implementation:

> *“…Whatever the government does, whether it’s the framework, the strategy, or the mechanisms, there must be transparency and accountability. That’s the number one priority. People need to be informed and sensitised so they can make informed decisions.”* (KII0001).

#### 3b. Strengthening Public Communication and Education

Stakeholders stressed that an effective pandemic response depends not only on securing vaccines but also on ensuring widespread public understanding and acceptance. They emphasised that without sustained education, even the best supply strategies would falter because misinformation, fear, and uncertainty could undermine uptake. As one district health director explained, awareness-building is critical to overcoming hesitancy:

> *“What is key is that, even if the country is able to procure the vaccines and the education does not go well, people will not take the vaccine. Thus, there is a need for community education.“* (KII0009).

Participants highlighted the importance of reaching every segment of society, stressing that education should continue until even the most remote communities understand the benefits of vaccination. These campaigns should go beyond basic information, providing clear and convincing messages about the advantages of vaccination to foster public trust:

> *“We need a lot of education for the last person to understand why they should be vaccinated, not just go and vaccinate for COVID-19; this way, we will all be fine. Yes, it is the education and then the district assemblies should provide more help, especially with the NCCE…* (KII0008).
>
> *“I think that there should be at least enough campaign and education of the public to know what is best, what the advantages of taking the vaccines are.”* (KII0003).

To achieve this, participants pointed to the need for stronger institutional support, particularly for the NCCE. They argued that education campaigns must be well-funded, consistent, and accessible across platforms. As one political actor put it:

> *“The NCCE should be properly funded because education of our people is very important for such exercises. So, there should be an advert on the radio so people become aware of what is going on.”* (KII0013)

#### 3c. Building Local Research and Vaccine Production Capacity

Stakeholders highlighted the importance of developing Ghana’s capacity to conduct research and manufacture vaccines locally, arguing that dependence on external suppliers had left the country vulnerable during the COVID-19 pandemic. They explained that procurement alone was not sufficient, as global shortages and inequities often limited access, underscoring the need for domestic capabilities. One pharmaceutical stakeholder described the vision of transforming Ghana into a regional hub for vaccine innovation and production:

> *“I think that we should be able to put together for the national immunity course and development, …but to also develop the ability to produce, the ability to make ourselves ready to be able to participate in global research to advance knowledge in health areas, and medical interventions like vaccines, to be able to also produce it, to be able to become a global hub even if we start from the West Africa…”* (KII0005).

Others affirmed this call, stressing that investment in research and development was critical to ensure timely access and reduce reliance on international partners in future crises:

> *“…that in future we would be able to invest a little further so that we can have our vaccines manufactured on time… Invest in research and development to make sure that some of these things are available to us and then ensure that distribution is more optimal”* (KII0006).

#### 3d. Enhancing Intersectoral Coordination and Leveraging Existing Structures

Stakeholders stressed the importance of improved coordination among ministries, agencies, the private sector and global partners to streamline future pandemic responses and prevent delays in vaccine rollout. They observed that limited integration of private providers during COVID-19 had undermined efficiency, despite their significant role in healthcare delivery. As one participant noted, excluding private facilities weakened national preparedness:

> “*The private health sector should be clearly included in government policy for interventions; otherwise, if the segregation continues, it will not augur well. There would be a dichotomy, and when the government needs them, it becomes difficult to rally them. I think 40% or more of healthcare facilities are in the private sector. And that is a significant number to be included in the planning for any pandemic…*(KII0014)

Others reinforced the need for more collaborative procurement strategies that allow both government and private institutions to contribute resources. A participant from academia suggested that shared efforts could improve speed and sustainability:

> “*I think another strategy is for us to see a more cohesive way of trying to procure the vaccine, both by the private and government institutions, so that the national COVID committee would assess the viability of this. It is quicker when more hands come on board. Because some of them, like the private sector, might have a way of financing and procuring these vaccines.*” (KII0011).

In addition to stronger intersectoral coordination, participants urged the government to leverage existing structures and expertise that had proven effective in past vaccination campaigns. Community health nurses and the EPI were highlighted as vital resources for scaling up future responses:

> “*We have a lot of community health nurses in the system. This is not the first time we are doing a vaccination exercise. … the system we are using has always been effective. When we do measles, the five childhood diseases…, we continue to do it perfectly. We have a system that moves to the remotest part of the country, where people would go on bicycles and stuff. We have a system, it exists already, so let’s use it*.” (KII0004).

Finally, participants highlighted the need to integrate trusted voices such as religious and traditional leaders to enhance vaccine acceptance. They argued that such leaders were instrumental in shaping community perceptions and could play a decisive role in mobilising participation:

> *“… so, I think they should talk to the people, and the churches can play a lot of roles, the mosques can play a major role too, they should use the pastors, they should use the Imams and get their people vaccinated”* (KII0007).
>
> “*Government should work with religious and traditional leaders as well as the influential people in society for the vaccination exercise.”* (KII0014)

#### 3e. Robust Monitoring and Evaluation Systems

Finally, participants underscored that future pandemic responses must be anchored in consistent monitoring, supervision, and evaluation to ensure accountability and effectiveness. They noted that while COVID-19 vaccination efforts mobilised significant resources, weak oversight limited the ability to track progress, identify bottlenecks, and measure impact. They argued that clear frameworks for monitoring would strengthen transparency and also provide timely feedback for corrective action. Some participants affirmed the need for supervisory teams capable of ensuring compliance and accountability across actors involved in the vaccination programme:

> *“…there should be monitoring and evaluation to be able to find out the impact of these policy measures”* (KII0006).
>
> “…*there should be a clear implementation plan and monitoring*” (KII0005).
>
> “*There should be a monitoring or supervisory team, tracking and checking these stakeholders*” (KII0010).

## Discussion

This study examined stakeholders’ perspectives on the political dynamics shaping Ghana’s COVID-19 mass vaccination programme, using the Walt & Gilson policy triangle and Campos & Reich’s policy implementation framework. The findings show that vaccine rollout outcomes were shaped less by technical capacity alone than by governance quality, institutional fragmentation, and global political economy constraints.

Transparency gaps, discretionary procurement practices, and weak digital oversight undermined trust and contributed to hesitancy, illustrating how accountability functions as a determinant of public health effectiveness. At the same time, political leadership, bureaucratic dynamics, budgetary prioritisation, donor dependence, and vaccine nationalism constrained procurement autonomy and distribution equity. Interest-group influence and socially embedded authority structures further mediated vaccine uptake.

Stakeholders’ proposed reforms, transparent governance, sustained public communication, domestic research and manufacturing capacity, stronger intersectoral coordination, and institutionalised monitoring collectively signal that pandemic preparedness must be treated as a governance and political economy challenge rather than a purely biomedical one. Strengthening future responses in Ghana and similar LMICs, therefore, requires embedding accountability, coordination, and adaptive governance within health security frameworks.

### Governance, Transparency, and Political Economy in Vaccine Rollout

Ghana’s early access to COVID-19 vaccines illustrates how high-level political commitment can accelerate entry into global supply chains. However, our findings demonstrate that political commitment alone does not guarantee effective implementation. Rather, vaccine rollout unfolded within a broader political economy of health governance characterised by transparency deficits, institutional fragmentation, donor dependence, and uneven operational capacity (30,31).

Participants’ concerns about opaque procurement and financial accountability reflect structural governance vulnerabilities widely documented in LMIC health systems (32). Political economy scholarship emphasises that emergency contexts expand discretionary authority while compressing oversight mechanisms, thereby increasing the risk of rent-seeking and elite capture (33). As Rose-Ackerman argues, weak transparency and oversight intensify corruption risks, particularly during crisis procurement (34). Similarly, Duri notes that pandemic emergencies create ideal conditions for financial opacity due to urgency, large inflows of funds, and relaxed procurement safeguards (35).

In this study, even the perception of politicised procurement eroded programme legitimacy. This aligns with broader LMIC evidence demonstrating that trust in vaccine programmes depends not only on technical efficiency but also on perceived fairness and integrity (36,37). Where procurement criteria and fund disbursement processes are unclear, suspicion can depress demand, strain inter-agency collaboration, and complicate donor engagement (38,39). Thus, transparency in vaccine rollout functions not merely as an ethical principle but as a core operational determinant of uptake and sustainability (40–42). Moreover, the blurred delineation of roles between central and subnational actors reflects classic principal-agent problems in health governance (4,43). When frontline implementers lack visibility into resource flows or decision criteria, implementation discretion increases and policy coherence weakens (4,44). These findings reinforce political economy arguments that governance quality, rather than supply availability alone, determines the success of emergency health interventions (45–47).

The communication failures described by participants highlight how vaccine uptake is embedded within broader dynamics of trust and authority. Delayed and fragmented risk communication allowed misinformation to circulate in both digital and religious spaces, shaping public perceptions before official narratives were consolidated (48–50). Global evidence shows that vaccine hesitancy during COVID-19 was strongly associated with distrust in government institutions, exposure to misinformation on social media, and reliance on informal authority figures (51,52). In many LMIC contexts, legitimacy is socially embedded: religious leaders, community elders, and informal opinion leaders often carry greater influence than technocratic institutions (32). The prominence of religious actors in shaping vaccine perceptions in Ghana reflects this broader pattern (50,52,53).

Importantly, misinformation should not be interpreted solely as a communication failure but as a political phenomenon (45,48). Where transparency deficits exist, conspiracy narratives find fertile ground (54,55). Political economy scholarship suggests that public compliance with health measures depends on institutional credibility and perceived procedural justice (45,46). In this regard, communication gaps interacted with governance weaknesses, reinforcing mistrust and slowing early uptake.

Ghana’s reliance on COVAX and bilateral donations situates the vaccine rollout within the wider geopolitical landscape of vaccine nationalism (12,13). The prioritisation of domestic populations by high-income countries, export restrictions, and production concentration in the Global North exposed structural asymmetries in global health governance (56–58). Political economy analyses of global vaccine distribution describe a core–periphery dynamic in which manufacturing capacity, bargaining power, and supply control are concentrated among wealthy states (30,59,60). Although COVAX aimed to mitigate inequities, supply unpredictability and delivery delays revealed the limits of multilateral solidarity in the face of national interest (61–63).

Our findings show how these global dynamics translated into local disruptions, missed second doses, scheduling uncertainty, and public frustration. This underscores a key insight that national vaccine rollout performance cannot be fully understood without reference to global political structures (59). While Ghana could not resolve global inequities domestically, buffering strategies, such as diversifying suppliers, securing predictable delivery windows, and maintaining transparent communication about supply constraints, could mitigate downstream effects (64). Participants’ scepticism regarding repeated announcements of local vaccine manufacturing further reflects broader debates about structural dependency (64). Without sustained institutional investment, domestic production remains aspirational, reinforcing long-standing reliance on external actors in health commodity supply chains.

The challenges observed in Ghana’s digital vaccination systems mirror a wider body of LMIC literature on electronic health information systems (64). While digital platforms are frequently promoted as tools for improving accountability and real-time monitoring, their effectiveness depends on infrastructural readiness, stable connectivity, trained personnel, and sustained financing (65–67). Studies from the WHO African Region and other LMIC settings consistently identify unreliable internet coverage, limited electronic devices, inadequate training, fragmented reporting systems, and delayed incentive payments as common barriers to digital health implementation (68–70). In this study, frontline reliance on manual workarounds and re-entry processes reflects these systemic constraints rather than isolated technical glitches (68). Importantly, digitalisation during crisis periods often outpaces institutional preparedness (71,72). Political incentives to demonstrate technological modernisation may result in the rapid rollout of applications without adequate piloting or capacity building (73,74). As such, digital failures should not be understood merely as operational inefficiencies but as governance challenges rooted in resource allocation, planning, and institutional coordination (75–77).

Cold-chain limitations and uneven distribution further demonstrate that vaccine rollout is embedded within broader infrastructural capacity (78,79). The technical requirements of mRNA vaccines exposed structural constraints common across sub-Saharan Africa, where ultra-cold storage and last-mile logistics remain unevenly developed (78,80–83).

The partial adaptation of the Expanded Programme on Immunisation (EPI) to adult mass vaccination highlights the limits of path dependency in health systems (84,85). While EPI platforms are robust for childhood immunisation, scaling to high-volume adult vaccination required different outreach strategies, communication models, and service delivery points (84,86).

Similarly, the limited integration of private pharmacies and facilities represents a missed opportunity in a mixed health system where a significant proportion of care is delivered outside the public sector (87). Political economy scholarship on health systems emphasises that exclusion of private actors can generate inefficiencies and coverage gaps, particularly in urban and peri-urban settings (88).

These findings demonstrate that vaccine rollout performance is shaped by layered political economy dynamics operating across global, national, and subnational levels (89). Governance transparency, institutional coordination, global power asymmetries, digital capacity, and social trust interact to determine policy outcomes.

Rather than viewing vaccine rollout challenges as isolated implementation failures, they should be understood as manifestations of structural governance and institutional capacity constraints common across LMICs (90). Strengthening preparedness for future outbreaks, whether influenza, mpox, or novel pathogens, requires treating governance, transparency, and institutional capacity as first-order operational investments rather than background administrative principles (91).

### Political Economy of Vaccine Procurement and Distribution

The findings demonstrate that vaccine procurement and distribution in Ghana were not merely technical public health exercises but politically negotiated processes embedded within domestic governance structures and global power asymmetries (64). Consistent with Campos and Reich’s political mapping framework, multiple actors, political elites, bureaucratic institutions, donors, and non-state stakeholders, shaped implementation through overlapping authority, competing incentives, and resource control. The centralisation of procurement authority and limited transparency in pricing negotiations reflect broader patterns identified in political economy analyses of emergency health governance in LMICs (92). Crisis procurement often concentrates decision-making power within executive structures while relaxing oversight mechanisms in the interest of speed (93). While such centralisation can accelerate action, it simultaneously increases discretion and reduces accountability safeguards (34).

Participants’ concerns regarding intermediaries and opaque pricing processes align with scholarship on rent-seeking in emergency procurement (94). Political economy theory suggests that when procurement systems lack transparency and institutional checks, opportunities for patronage, preferential contracting, or perceived elite capture increase, even where formal corruption cannot be established (93). Importantly, in vaccine rollouts, perception itself becomes politically consequential: suspicion of preferential allocation or inflated pricing can undermine public trust and reduce compliance with vaccination campaigns (30).

The reported extension of political authority into subnational distribution further reflects how decentralised implementation remains vulnerable to partisan (95). Even isolated perceptions of political gatekeeping at district levels illustrate how public health programmes may become entangled in local patronage networks (96). This aligns with broader governance literature in sub-Saharan Africa, demonstrating that distributive goods, including health commodities, can be politicised where accountability mechanisms are weak and electoral competition is salient (97,98).

Bureaucratic friction between policy design and operational execution also reflects structural institutional fragmentation (99,100). Health systems in many LMICs operate through layered hierarchies in which ministries set policy while semi-autonomous agencies manage implementation (47,101). Where role clarity and coordination mechanisms are insufficient, implementation gaps emerge despite strong planning processes (102). The partial realisation of planned pharmacy integration illustrates how bureaucratic discretion and institutional inertia can dilute reform ambitions (102).

Budgetary politics further compounded these governance challenges. Although procurement financing was prioritised, delayed or insufficient funding for deployment reveals how fiscal allocation reflects political prioritisation rather than purely epidemiological need (103,104). Political economy scholarship emphasises that budgets are expressions of power; allocation decisions determine which aspects of policy are realised and which remain aspirational (47,103). In this case, deployment constraints, particularly in rural districts, demonstrate how fiscal bottlenecks translate into geographic inequities in access (47,102,103).

Ghana’s dependence on donor-supported supply chains situates the vaccine rollout within the broader geopolitical phenomenon of vaccine nationalism (47). As documented globally, high-income countries secured advance purchase agreements that limited early access for countries reliant on pooled mechanisms such as COVAX (105). Political economy analyses characterise this dynamic as a manifestation of structural inequality in global health governance (47).

While multilateral platforms aimed to mitigate inequities, participants’ accounts reflect how global supply unpredictability produced local implementation instability (63). Delayed shipments, export restrictions during domestic surges, and fluctuating delivery timelines illustrate the vulnerability of peripheral economies within global commodity chains (47,63). These findings align with scholarship arguing that COVAX, although normatively grounded in equity, operated within an international system still governed by national self-interest (47,63).

Moreover, donor dependence constrained national autonomy in procurement planning and scheduling (106). Political economy theory suggests that aid-dependent sectors often experience reduced policy space, as timelines and availability are shaped externally (47,103). The resulting uncertainty not only disrupted vaccination cycles but also reinforced perceptions of structural vulnerability, fuelling domestic debates about sovereignty and self-reliance.

Participants’ emphasis on local vaccine manufacturing, therefore, reflects more than economic aspiration; it signals recognition of structural dependency within global pharmaceutical production networks (47). Without sustained institutional investment, however, such ambitions risk remaining rhetorical rather than transformative (47,107).

The findings also reveal how non-state actors influenced both uptake and strategic direction (108). Professional associations advocating for domestic production illustrate how interest groups seek to shape long-term supply architecture through policy influence (47,107). Their demands for procurement guarantee reflect classic industrial policy debates, where state purchasing commitments are used to stimulate local manufacturing capacity (47,103,107).

Religious leaders’ influence on vaccine decisions underscores the socially embedded nature of authority in many LMIC contexts (109). Political economy scholarship on public health compliance highlights that trust in government institutions is often mediated by intermediary actors (109). Where institutional trust is uneven, community members may rely more heavily on faith leaders, community figures, or informal networks when making health decisions (47,109).

The reported influence of religious authority, even among medically trained professionals, demonstrates how legitimacy operates through social identity and moral authority rather than technical expertise alone (47,109). In such contexts, vaccine acceptance cannot be framed solely as an information deficit problem. Instead, it reflects negotiated trust relationships between state institutions and socially embedded power structures.

At the same time, doctrinal resistance within certain communities illustrates how health policy can intersect with ideological worldviews (109–111). Political economy approaches emphasise that health interventions are interpreted within cultural and political narrative, where governance transparency is weak, scepticism toward state initiatives may become entangled with religious or political identities (109,110).

These findings illustrate how vaccine procurement and distribution were shaped by multi-level political dynamics. Rather than viewing vaccine rollout challenges as isolated administrative shortcomings, they should be understood as outcomes of interacting political economy forces operating across scales (105,110). Effective pandemic preparedness, therefore, requires institutional reforms that address transparency, fiscal prioritisation, bureaucratic coordination, and structured engagement with influential non-state actors (105).

### Lessons for Future Pandemics

The lessons identified by stakeholders reveal deeper structural insights into how pandemic preparedness is shaped by governance quality, institutional capacity, and global political economy dynamics (63). Rather than viewing these lessons as technical improvements, they should be interpreted as institutional reforms addressing systemic vulnerabilities exposed during COVID-19.

Participants’ emphasis on transparency and accountability reflects recognition that governance is not peripheral to pandemic response but constitutive of it (112). Political economy scholarship consistently demonstrates that where procurement systems lack transparency and oversight, public trust erodes and policy compliance weakens (34). In emergency settings, rapid fund mobilisation and discretionary authority increase corruption risks unless counterbalanced by institutional safeguards (113). Calls for independent oversight structures and publicly accessible reporting mechanisms reflect broader governance reform debates in LMIC health systems (114,115). Transparency reduces informational asymmetries between state actors and citizens, thereby limiting opportunities for rent-seeking and strengthening legitimacy (116). Importantly, legitimacy itself functions as a public health asset: where community members perceive fairness and procedural justice, compliance with vaccination campaigns improves (117–119). Thus, transparent governance is not merely an ethical aspiration but a functional prerequisite for effective outbreak response.

The strong emphasis on public communication underscores that vaccine uptake is embedded within social trust dynamics rather than solely determined by supply availability (120–122). Political economy and public health compliance literature emphasise that effective risk communication must be institutionalised before crises occur (123,124). Reactive communication strategies, deployed only after misinformation has proliferated, struggle to rebuild eroded trust (118).

The recommendation to strengthen the National Commission for Civic Education (NCCE) and leverage religious and community leaders reflects an understanding of socially distributed authority in LMIC contexts (52). In many societies, trust is mediated through relational networks rather than formal state institutions (52). Embedding these actors within preparedness frameworks acknowledges that health behaviour is shaped by political legitimacy, moral authority, and social identity (52). These findings align with global evidence that vaccine hesitancy often reflects institutional distrust rather than informational deficits (118,125,126). Therefore, communication reform should be conceptualised not as messaging enhancement alone, but as a broader strategy for rebuilding state–citizen trust relationships (118,126,127).

Participants’ advocacy for domestic research and vaccine manufacturing capacity speaks directly to structural dependency within global pharmaceutical supply chains (128,129). The COVID-19 pandemic revealed that LMIC reliance on external production subjects national health security to geopolitical bargaining and supply nationalism (130–132). Political economy analyses describe this as asymmetrical interdependence: peripheral economies depend on core manufacturing hubs with limited reciprocal leverage (128,129). Calls for local production. therefore, reflect more than economic ambition; they represent an attempt to expand national policy autonomy within global health governance structures (133). However, scholarship on industrial policy in LMICs cautions that local production requires sustained capital investment, technology transfer agreements, regulatory strengthening, and guaranteed procurement mechanisms (128,129,133). Without institutional continuity and political commitment beyond crisis periods, manufacturing aspirations risk remaining symbolic (128,129,133). The findings thus reinforce the argument that pandemic preparedness must incorporate industrial policy considerations, not merely epidemiological planning (128). Vaccine self-reliance, even at regional scale, functions as a strategic hedge against global supply volatility (128).

Stakeholders’ emphasis on stronger intersectoral coordination reflects recognition of fragmentation within health governance systems (6). Political economy research highlights that crisis responses often create parallel structures that bypass existing institutions, generating duplication and inefficiency (6). Integrating private providers, community health structures, and established immunisation platforms such as EPI into preparedness planning represents an institutional consolidation strategy rather than simple service expansion (4).

The limited engagement of private sector actors during COVID-19 illustrates the cost of excluding mixed health system components (4,6). In many LMICs, significant portions of primary care are delivered outside the public sector; excluding these actors reduces surge capacity and geographic reach (4,6). Whole-of-society approaches, frequently advocated in global health governance frameworks, require formalised coordination mechanisms rather than ad hoc collaboration.

Leveraging existing structures, community health nurses, established immunisation outreach systems, and faith-based networks, aligns with resilience theory in health systems (6). Strengthening preparedness, therefore, requires institutional alignment across sectors before emergencies emerge.

Participants’ call for robust monitoring and evaluation systems reflects the need for adaptive governance during health crises (5,6). Political economy scholarship underscores that emergency responses are dynamic; policies must evolve in response to emerging evidence, operational bottlenecks, and shifting public sentiment. Without structured monitoring frameworks, governments lose the capacity for course correction (4–6). Embedding monitoring mechanisms into preparedness frameworks enhances transparency, reduces implementation drift, and institutionalises learning (4–6). Adaptive governance models emphasise iterative feedback loops, enabling policy refinement rather than rigid adherence to initial plans (4–6). In LMIC contexts where data systems may be fragmented, strengthening monitoring capacity also mitigates informational asymmetries (4–6).

Investment in digital health infrastructure, when accompanied by institutional readiness and oversight, can strengthen surveillance and coordination (4,134). However, as demonstrated in broader LMIC literature, digital tools must be embedded within governance reforms to avoid replicating existing accountability gaps (134). These lessons demonstrate that pandemic preparedness cannot be reduced to vaccine stockpiling or biomedical readiness. Instead, preparedness is a function of governance quality, institutional coherence, fiscal prioritisation, global positioning, and social trust (4).

For Ghana and comparable LMICs, embedding transparency, strengthening institutional coordination, investing in domestic production capacity, formalising engagement with socially legitimate actors, and institutionalising adaptive monitoring mechanisms represent structural reforms rather than temporary adjustments. Political and governance dynamics ultimately determine whether technical interventions translate into equitable and effective vaccine delivery (4,5).

Future outbreaks, whether influenza, mpox, or novel pathogens, will test not only health systems but also governance systems. Pandemic resilience, therefore, depends on treating political economy reform as central to health security strategy rather than as a secondary consideration (4).

### Limitations

Not all intended stakeholder groups were represented, as international donor agencies, some religious leaders, political actors, and civil society coalitions did not take part in the study, which may have limited the breadth of perspectives captured. The sample size was relatively small (14 interviews) and largely drawn from Greater Accra, which limits the breadth of regional insights. Additionally, data were collected in 2021, which reflects only the early implementation and not subsequent developments. As a qualitative study, findings are not statistically generalisable but provide contextual insights into governance, politics and communication. Despite these limitations, the study offers important lessons for strengthening accountability, communication and future preparedness in future health emergencies.

## Conclusion

This study demonstrated how political dynamics shaped Ghana’s COVID-19 vaccination programme. While early and strong political commitment facilitated vaccine access, gaps in coordination, limited accountability and transparency hindered effective implementation. Procurement and distribution were further complicated by financial constraints, donor politics, and influence of interest groups and beneficiaries, highlighting that vaccine delivery is as much a political process as it is a technical one. Transparent and accountable governance, timely financing, effective stakeholder coordination and robust public communication are vital in sustaining trust and ensuring equitable access. Strengthening Ghana’s capacity for local vaccine, research and production, alongside investment in monitoring and evaluation, will be very important for reducing dependence on external actors and safeguarding future security. Although focused on Ghana, these findings are highly relevant across LMICs facing a similar structure and political challenges. As global health systems prepare for future health threats, effective governance, political alignment and communities determine whether life-saving technologies reach those who need them most.

## Key Findings

- The government’s strong leadership in initiating a nationwide COVID-19 vaccination programme demonstrates the nation’s commitment to pandemic control, despite significant implementation challenges such as coordination gaps, limited transparency, and weak accountability.
- Procurement and distribution were hindered by political leadership, bureaucratic processes, budget constraints, donor politics, and the influence of interest groups and beneficiaries.
- Misinformation and vaccine hesitancy, amplified by social media spreading false information, slowed uptake and underscored the importance of trusted communication.
- Stakeholders underscored the need for transparency, strong monitoring and evaluation, and investment in local research and vaccine production to strengthen preparedness for future health emergencies.

## Key Implications for

- Strengthen governance and accountability structures to ensure transparent procurement and resource allocation, and invest in local vaccine research and production to reduce external dependence.
- Coordinated frameworks should be built across ministries and agencies, timely disbursement of resources to frontline actors ensured, and culturally grounded, community-driven communication strategies developed to counter misinformation and reduce hesitancy.
- Building regional manufacturing capacity and equitable allocation mechanisms to reduce low- and middle-income countries’ vulnerability to external supply disruptions.

## Acknowledgements

We would like to express profound gratitude to all study participants and data transcribers whose contributions and efforts were indispensable to the integrity and success of this research.

## Data Availability

The qualitative interview data generated for this study are not publicly available due to ethical and legal restrictions related to participant confidentiality. Although transcripts were de-identified, the specific roles of participants and the sensitive policy context create a risk of deductive disclosure. Data may be made available to qualified researchers upon reasonable request and with approval from the ethics oversight body. Requests should be directed to the Dodowa Health Research Centre Institutional Review Board, Dodowa, Ghana, and copied to the corresponding author.

## Authors Contributions

DEA conceptualized the study, contributed to the proposal development, conducted interviews, and supported data analysis and report writing. RAN was involved in the proposal writing, coding, data analysis, and interpretation of findings. RH contributed substantially to the drafting of the initial manuscript and provided critical review of the research report. MI provided overall supervision, guided the manuscript development, and reviewed all versions of the research report and final manuscript. All authors reviewed, revised, and approved the final version of the manuscript for submission.

